# The Mediterranean Diet, Cardiometabolic Biomarkers, and Risk of All-Cause Mortality: A 25-Year Follow-Up Study of the Women’s Health Study

**DOI:** 10.1101/2023.10.02.23296458

**Authors:** Shafqat Ahmad, M. Vinayaga Moorthy, I-Min Lee, Paul M Ridker, JoAnn E. Manson, Julie Buring, Olga V. Demler, Samia Mora

## Abstract

**Background:** Higher consumption of Mediterranean diet (MED) intake has been associated with reduced risk of all-cause mortality but limited data are available examining long-term outcomes in women or the underlying molecular mechanisms of this inverse association in human populations. We aimed to investigate the association of MED intake with long-term risk of all-cause mortality in women and to better characterize the relative contribution of traditional and novel cardiometabolic factors to the MED-related risk reduction in morality.

**Methods:** In a prospective cohort study of 25,315 initially healthy women from the Women’s Health Study, we assessed dietary MED intake using a validated semiquantitative food frequency questionnaire according to the usual 9-category measure of MED adherence. Baseline levels of more than thirty cardiometabolic biomarkers were measured using standard assays and targeted nuclear magnetic resonance spectroscopy, including lipids, lipoproteins, apolipoproteins, inflammation, glucose metabolism and insulin resistance, branched-chain amino acids, small metabolites, and clinical factors. Mortality and cause of death was ascertained prospectively through medical and death records.

**Results:** During a mean follow-up of 25 years, 3,879 deaths were ascertained. Compared to the reference group of low MED intake (0-3, approximately the bottom tertile), and adjusting for age, treatment, and energy intake, risk reductions were observed for the middle and upper MED groups with respective HRs of 0.84 (95% CI 0.78-0.90) and 0.77 (95% CI 0.70-0.84), p for trend <0.0001. Further adjusting for smoking, physical activity, alcohol intake and menopausal factors attenuated the risk reductions which remained significant with respective HRs of 0.92 (95% CI 0.85-0.99) and 0.89 (95% CI 0.82-0.98), p for trend 0.0011. Risk reductions were generally similar for CVD and non-CVD mortality. Small molecule metabolites (e.g., alanine and homocysteine) and inflammation made the largest contributions to lower mortality risk (accounting for 14.8% and 13.0% of the benefit of the MED-mortality association, respectively), followed by triglyceride-rich lipoproteins (10.2%), adiposity (10.2%) and insulin resistance (7.4%), with lesser contributions (<3%) from other pathways including branched-chain amino acids, high-density lipoproteins, low-density lipoproteins, glycemic measures, and hypertension.

**Conclusions:** In the large-scale prospective Women’s Health Study of 25,315 initially healthy US women followed for 25 years, higher MED intake was associated with approximately one fifth relative risk reduction in mortality. The inverse association was only partially explained by known novel and traditional cardiometabolic factors.

## INTRODUCTION

Nutrition and prevention guidelines focus on adherence to dietary patterns rather than single food nutrients in relation to health outcomes^1–6^. A recent umbrella review of 495 unique meta-analyses of observational studies and randomized clinical trials examined the associations with a wide range of different dietary patterns in relation to cardiometabolic and anthropometric risk factors. The review reported that the Mediterranean diet (MED) had the most consistent associations with lower risks of chronic diseases^7^. The US dietary guidelines have repeatedly designated the MED diet as the healthiest recommended diet^8^. Guidelines from the American Heart Association, European Society of Cardiology and Australian National Heart Foundation have consistently highlighted MED as a healthy dietary pattern for improving cardiometabolic health and cardiovascular disease (CVD) outcomes^9–11^.

Many large-scale observational epidemiological studies with long follow-up support an association between higher adherence to MED and reduced risk of all-cause mortality^3, 12–19^. A meta-analysis based on 29 observational studies which included 1,676,901 participants reported that a 2-point increase in the consumption of MED was associated with 10% reduction in all-cause mortality^20^, while another meta-analysis of 21 cohort studies which included 883,878 participants reported that higher MED intake was associated with 21% reduced risk of CVD mortality^21^. However, long-term all-cause mortality data in asymptomatic women are limited. Hence, we aimed to investigate the association of MED intake with long-term risk of all-cause mortality in women, Furthermore, the precise mechanisms through which increased adherence to the MED diet associates with lower risk of mortality are poorly understood, especially the relative contribution of traditional and newly discovered novel cardiometabolic biomarkers related to inflammation, lipids and lipoproteins, glucose metabolism and insulin resistance, branched-chain amino acids (BCAAs). A recent systematic review has reported that higher MED intake was associated with improved inflammatory biomarkers including C-reactive protein (CRP)^22^. The Prevención con Dieta Mediterránea (PREDIMED) randomized trial conducted in Spain where participants were followed-up for 4-years reported that MED intake was associated with increased large high-density lipoprotein (HDL) particles, significantly reduced low-density lipoprotein (LDL) levels and diastolic blood pressure,^23^ which has also been seen in observational studies ^24–26^ but the relative contribution of these pathways to lower mortality risk is unknown. Additionally, prior studies did not incorporate new biomarkers representing additional cardiometabolic pathways.

Therefore, in a large-scale prospective epidemiological cohort study of 25,315 initially healthy women from the US population with 25-year follow-up, we aimed to investigate whether higher adherence to MED was associated with lower long-term risk of mortality and to quantify the contribution of traditional as well as novel biological biomarkers to the MED associated mortality reduction.

## MATERIALS AND METHODS

### Study population

Participants were enrolled in the prospective Women’s Health Study (WHS), as described in detail previously^27–29^. Briefly, a total of 39,876 healthy healthcare female professionals aged 45 years at baseline (1992-1995) were randomized into low dose aspirin and Vitamin E or their corresponding placebos. On the baseline questionnaire, participants provided information about demographics, anthropometry, lifestyle, medical and social history, and medications. They also provided self-reported weight and height and body mass index (BMI) was calculated. Diastolic and systolic blood pressures were reported at baseline. All study participants provided written consent and the study was approved by the institutional review board of Brigham and Women’s Hospital, Boston, Massachusetts.

For the current analysis, we included 28,340 participants who provided blood samples at baseline, then excluded participants who do not have biomarkers measurements as well as dietary information, leaving a total sample of n=25,315 for the current study.

### Mediterranean Diet (MED) Intake

For the assessment of dietary patterns, a validated semiquantitative food frequency questionnaire of 131 questions was administered to the study participants at baseline^30^. The MED score has been previously described in detail^31^. Briefly, the MED score ranged from 0 to 9 where a higher score represents better adherence to MED. This MED score is commonly used for assessing adherence to MED diet, and is based on regular intake of nine dietary components where higher than median intake of vegetables (excluding potatoes), fruits, nuts, who-grains, legumes, fish, ratio to monounsaturated-to-saturated fatty acids were given 1 point while the intake of red and processed meat intake was given 1 point if the intake was less than the median intake. For constructing the MED score, participants were given 1 point if the intake of medium alcohol consumption was from 5 to 15 g/d (otherwise 0 points). Participants were categorized into three levels based on MED intake: scores of 0-3, 4-5 and 6-9^32^, representing approximate tertiles.

### Mortality Ascertainment

Detailed description of mortality ascertainment in the WHS cohort has been described in detail^33, 34^. In brief for all cases, death records were obtained and reviewed by an Endpoints Committee. Death information was collected through the National Death Index, and death certificates including circumstances of death. Cause-specific mortality including CVD deaths were ascertained only based on death certificates and end point committee adjudication to limit potential for misclassification. CVD mortality included deaths caused by ischemic heart, acute myocardial infarction, cerebrovascular, sudden death and other cardiovascular-related deaths.

### Blood Collection and Measurement of Traditional Biomarkers

At baseline, participants’ blood samples were collected in EDTA tubes and shipped overnight to the central laboratory where they were centrifuged and stored at -170°C until further analyses. The concentrations of hemoglobin A1c in red blood cells was measured through turbidometric assays through Hitachi 911 Analyzer (Roche Diagnostics, Indianapolis, Ind). The measurement of high-sensitivity C-reactive protein (hsCRP) and lipoprotein(a) (Lp[a]) was performed by turbidometric immunoassays through Hitachi-911 analyzer (Roche Diagnostics)^28^. The measurement of total cholesterol, LDL cholesterol (LDL-C), HDL-C were enzymatically performed using assays from Roche Diagnostics and Genzyme. Triglycerides (TG) was measured through enzymatic assays based on Roche Diagnostics after correction for endogenous glycerol. Apolipoproteins (apo) A1 and B100 were measured using turbidometric assays (DiaSorin). The measurement of fibrinogen was performed using turbidometric enzymatic assay (R&D Systems). The measurement of soluble intracellular adhesion molecule 1 (ICAM-1) was enzymatically performed through immunosorbent assay (R&D Systems). The measurement of creatinine was performed using a rate-blanked Jaffe reaction-based method (Roche Diagnostics). The measurement of homocysteine was enzymatically performed using the Hitachi-917 analyzer (Roche Diagnostics) through using the reagents and calibrators from the Catch, Inc.

### Nuclear magnetic resonance spectroscopy metabolomics biomarkers

Targeted nuclear magnetic resonance (NMR) spectroscopy^31, 35, 36^ of ^1^H-NMR (400 MHz) LipoProfile-IV (LipoScience; now LabCorp) was used to measure lipoprotein subfraction particles of LDL, HDL, very low-density lipoprotein (VLDL)/triglyceride-rich lipoproteins (TRL), and small molecule metabolites. The NMR-based lipoprotein insulin resistance score (LPIR) which includes subfractions of TRL, LDL and HDL particles^37^. The NMR assay was also used to measure glycoprotein acetylation (GlycA, an aggregate inflammatory biomarker of circulating glycosylated acute phase proteins), as well as other cardiometabolic small molecules including alanine, citrate, branched-chain amino acids (BCAA [leucine, isoleucine, valine]) and others. Insulin resistance index which is a multimarker score which combines lipoprotein insulin resistance and 5-years diabetes risk factor index were also measured using NMR assay.

## STASTICAL ANALYSES

We used Cox proportional hazards regression models to compute adjusted hazards ratio (HRs) for mortality as well as their corresponding 95% CIs through employing MED intake (0-3, approximately bottom tertile) as a reference category. To compute the linear trend estimates, we used the median values of the three MED intake categories (0-3, 4-5, 6-9). Two-sided *p*-values P value threshold >0.05 was used for the mediation analysis. The measures of TG, hsCRP, Lp(a) and homocysteine were not normally distributed and were log-transformed.

To test whether biomarkers fulfilled the criteria to be used as mediators, the Baron and Kenny approach was used^38^. Mediation analyses were performed through the standard mediation approach^39^, however, for single biomarkers as mediators we also computed a counter-factual framework approach along-with standard mediation approach to calculate total mediated effect^40^. One of the caveats of using counterfactual framework approach is that it cannot be applied to multiple biomarkers simultaneously. Person-years of follow-up were computed from baseline until death or censoring.

First, we tested the significance of association of MED intake with incidence of all-cause and CVD mortality. After that, through using a separate model for each of the potential biomarker, we tested again the association of MED intake with the risk of mortality. Statistical models were adjusted for age, randomized treatment assignment, and energy intake, and additionally in the base model for smoking, physical activity, and menopausal factors. Change in magnitude of HRs for the highest (>6) vs lowest (0-3) MED intake group by adjusting or not adjusting for additional variables one at a time. A greater change in the HR towards null means that a larger mediating effect through that particular biomarker on the MED intake-associated reduction in the mortality risk.

Further, on *an priori* hypothesis basis, we grouped biomarkers into specific groups based on the potential biological mechanisms. Details about biomarkers groups have previously been discussed in detail^41^. Then, we assessed the change in the magnitude of the HRs comparing lowest vs highest MED intake in association with the mortality incidence by adding one group at a time to the base model (adjusting for baseline age, randomization treatment assignment, energy intake, postmenopausal use of hormones, postmenopausal status, physical activity and smoking). To examine the extent through which each set of risk factors mediate the effect of MED intake in association of mortality risk, we added one group of biomarkers, at a time, to the basic models and compared the change in magnitude of the HRs for the group with highest intake in MED intake with the lowest without and with adjustment of each mediator set. A larger change in the HRs toward null implies a great change by the mediator group about the association of MED intake with mortality risk.

The proportion of mortality risk reduction explained by each group of biomarkers was inferred using the formula:

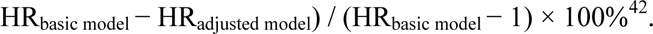

Statistical analyses were performed using Stata version 14.0 (StataCorp) and SAS version 9.3 (SAS Institute).

## RESULTS

### Baseline characteristics

Participants reporting higher adherence to MED intake had general healthier lifestyles, lower adiposity, higher consumption of fruits, nuts, whole grains, legumes and fish while lower intake of red and processed meat (**Table 1**). There were modest but statistically significant differences in all biomarker and risk factor profiles except for systolic blood pressure, LDL cholesterol, apolipoprotein B-100, LDL particle concentration, and creatinine. Higher MED intake was associated with an overall better biomarker profile (*P* for linear trend < 0.05).

**Table 1.** Baseline biomarker levels according to Mediterranean diet intake.

| Mediterranean Diet Intake |  |  |  |  |
| --- | --- | --- | --- | --- |
| Median (IQR) |  |  |  |  |
| Biomarker | MED Score 0-3 (n=9,871) | MED Score 4-5 (n= 9,184) | MED Score ≥ 6 (n=6,260) | P Value for Trend |
| Age, median (IQR), y | 51.9 (48.4-57.3) | 53.3 (49.1-59.1) | 54.1 (49.8-60.6) | <0.0001 |
| Current smoking, % | 15.7 | 10.4 | 6.3 | <0.0001 |
| Exercise, % |  |  |  |  |
| Rarely/Never | 44.7 | 35.4 | 26.0 | <0.0001 |
| <1 /week | 20.3 | 19.0 | 19.2 |  |
| 1-3/week | 27.0 | 33.7 | 38.5 |  |
| 4+ /week | 8.0 | 11.9 | 16.3 |  |
| Alcohol consumption, % |  |  |  |  |
| Rarely | 49.2 | 41.8 | 35.0 | <0.0001 |
| 1-3 drinks/month | 14.6 | 13.4 | 11.0 |  |
| 1–6 drinks/week | 27.8 | 33.8 | 40.3 |  |
| ≥1 drinks/day | 8.4 | 10.0 | 13.8 |  |
| Vegetable intake, median (IQR), servings/day | 2.3 (1.6-3.1) | 3.7 (2.8-5.0) | 5.2 (4.1-6.8) | <0.0001 |
| Fruits, median (IQR), servings/day | 1.3 (0.8-1.8) | 2.1 (1.4-2.9) | 2.8 (2.2-3.7) | <0.0001 |
| Nuts, median (IQR), servings/day | 0 (0-0.07) | 0.07 (0-0.13) | 0.07 (0-0.1) | <0.0001 |
| Whole grains, median (IQR), servings/day | 0.7 (0.3-1.1) | 1.2 (0.7-1.9) | 1.8 (1.3-2.8) | <0.0001 |
| Legumes, median (IQR), servings/day | 0.2 (0.1-0.4) | 0.4 (0.2-0.6) | 0.6 (0.4-0.9) | <0.0001 |
| Fish, median (IQR), servings/day | 0.1 (0.07-0.2) | 0.2 (0.1-0.3) | 0.3 (0.2-0.5) | <0.0001 |
| Ratio of monounsaturated to saturated fat | 1.1 (1.0-1.2) | 1.1 (1.0-1.2) | 1.2 (1.1-1.3) | <0.0001 |
| Red meat, median (IQR), servings/day | 0.6 (0.4-1.0) | 0.6 (0.3-1.0) | 0.5 (0.3-0.9) | <0.0001 |
| Processed meats, median (IQR), servings/day | 0.1 (0.07-0.3) | 0.1 (0-0.2) | 0.07 (0-0.2) | <0.0001 |
| Postmenopausal status, % | 49.8 | 55.3 | 59.0 | <0.0001 |
| **Adiposity** |  |  |  |  |
| BMI, kg/m<sup>2</sup> | 25.0 (22.5-28.3) | 24.8 (22.5-28.2) | 24.2 (22.1-27.4) | <0.0001 |
| **Blood pressure, mmHg** |  |  |  |  |
| Systolic | 125.0 (115.0-135.0) | 125.0 (115.0-135.0) | 125.0 (115.0-135.0) | 0.65 |
| Diastolic | 80.0 (70.0-80.0) | 80.0 (70.0-80.0) | 80.0 (70.0-80.0) | 0.095 |
| **Lipids, mg/dL** |  |  |  |  |
| LDL | 121.6 (100.7-144.4) | 121.1 (100.9-144.1) | 121.4 (100.3-144.7) | 0.91 |
| HDL | 51.5 (43.0-61.6) | 52.7 (43.9-63.1) | 53.8 (44.9-64.5) | <0.0001 |
| Triglycerides | 117.0 (83.0-171.0) | 117.0 (83.0-172.0) | 116.0 (83.0-167.0) | 0.15 |
| Total cholesterol | 207.0 (183.0-234.0) | 208.0 (184.0-235.0) | 208.0 (184.0-236.0) | 0.014 |
| **Lipoproteins, mg/dL** |  |  |  |  |
| Lipoprotein(a) | 10.4 (4.4-32.3) | 10.9 (4.7-33.5) | 10.8 (4.5-32.9) | 0.12 |
| Apolipoprotein A1 | 148.0 (131.3-166.5) | 150.0 (133.7-168.5) | 152.0 (135.3-171.0) | <0.0001 |
| Apolipoprotein B-100 | 99.8 (83.8-120.4) | 99.5 (83.2-120.1) | 99.2 (83.5-120.5) | 0.86 |
| **LDL particles and size** |  |  |  |  |
| LDL particle concentration, nmol/L | 1428.7 (1194.2-1699.1) | 1430.0 (1198.8-1695.7) | 1433.9 (1194.9-1696.8) | 0.99 |
| LDL particle size, (nm) | 21.0 (20.7-21.3) | 21.1 (20.7-21.3) | 21.1 (20.8-21.3) | 0.0001 |
| **HDL particles and size** |  |  |  |  |
| HDL particle concentration, μmol/L | 22.3 (20.2-24.5) | 22.5 (20.5-24.7) | 22.6 (20.6-24.9) | <0.0001 |
| HDL particle size, (nm) | 9.0 (8.7-9.3) | 9.0 (8.8-9.4) | 9.1 (8.8-9.4) | <0.0001 |
| **TRL particles and size** |  |  |  |  |
| TRL particle concentration, nmol/L | 163.0 (125.2-206.1) | 161.3 (122.9-205.6) | 160.8 (122.1-206.1) | 0.13 |
| TRL particle size, nm | 42.9 (38.7-48.5) | 42.8 (38.8-48.4) | 42.7 (38.8-48.0) | 0.23 |
| **Glycemic measures** |  |  |  |  |
| Hemoglobin A1c, % | 5.0 (4.8-5.2) | 5.0 (4.8-5.2) | 5.0 (4.8-5.2) | 0.013 |
| **Insulin resistance** |  |  |  |  |
| Lipoprotein insulin resistance index score | 45.0 (27.0-62.0) | 44.0 (26.0-61.0) | 42.0 (25.0-59.0) | <0.0001 |
| 5-y diabetes risk factor index score | 41.0 (28.0-55.0) | 40.0 (28.0-53.0) | 39.0 (28.0-52.0) | <0.0001 |
| **Inflammation** |  |  |  |  |
| High-sensitivity C-reactive protein, mg/L | 2.0 (0.8-4.4) | 2.0 (0.8-4.2) | 1.8 (0.7-3.9) | <0.0001 |
| Fibrinogen, mg/dL | 350.3 (306.7-403.3) | 349.7 (307.5-400.1) | 346.1 (304.6-396.7) | 0.0019 |
| Soluble intercellular adhesion molecule 1, ng/mL | 344.4 (301.5-397.5) | 341.2 (299.8-390.6) | 337.1 (297.3-383.5) | <0.0001 |
| Glycoprotein acetylation, μmol/L | 374.7 (330.9-422.4) | 372.6 (330.0-418.7) | 368.9 (326.1-413.6) | <0.0001 |
| **Branched-chain amino acids, μmol/L** |  |  |  |  |
| Total branched-chain amino acids | 382.2 (334.6-436.5) | 378.6 (333.6-431.7) | 376.5 (331.4-427.3) | <0.0001 |
| Valine | 190.1 (165.0-218.4) | 188.6 (165.2-215.6) | 187.6 (164.6-214.4) | 0.0042 |
| Leucine | 128.0 (107.4-150.3) | 126.6 (105.6-149.2) | 126.2 (106.3-148.3) | 0.001 |
| Isoleucine | 64.7 (53.8-77.3) | 64.0 (53.5-76.0) | 63.2 (53.2-74.7) | <0.0001 |
| **Small molecule metabolites** |  |  |  |  |
| Citrate, μmol/L | 99.0 (84.7-114.6) | 99.5 (84.3-115.8) | 98.2 (83.5-114.8) | 0.019 |
| Creatinine, mg/dL | 0.7 (0.6-0.8) | 0.7 (0.6-0.8) | 0.7 (0.6-0.8) | 0.33 |
| Homocysteine, μmol/L | 10.7 (8.8-13.2) | 10.3 (8.6-12.7) | 10.2 (8.6-12.4) | <0.0001 |
| Alanine, mg/dL | 3.8 (3.3-4.4) | 3.9 (3.4-4.5) | 4.0 (3.5-4.5) | <0.0001 |

Abbreviations: HDL, high-density lipoprotein; HR, hazard ratio; LDL, low-density lipoprotein; MED, Mediterranean diet; TRL, triglyceride-rich lipoprotein. SI conversion factors: To convert HDL and LDL cholesterol to mmol/L, multiply by 0.0253; triglycerides to mmol/L, multiply by 0.0113; lipoprotein(a) to $\mu$ mol/L, multiply by 0.0357; apolipoprotein AI and Apolipoprotein B100 to g/L, multiply by 0.01; hemoglobin A1c to proportion of total hemoglobin, multiply by 0.01; C-reactive protein to nmol/L, multiply by 9.524; fibrinogen to $\mu$ mol/L, multiply by 0.0294; and creatinine to $\mu$ mol/L, multiply by 88.4. aWe categorized the participants according to 3 levels of MED (scores of 0-3, 4-5, and 6-9). P values across 3 levels of MED were all less than .05.

Most of the biomarkers met Baron and Kenny’s criteria for mediation (**Table 1**), except for systolic blood pressure, LDL cholesterol, Apo B-100, LDL particle concentration, TRL particle concentration, hemoglobin A1c, and creatinine (all *P*>0.05). However, significant associations between higher MED intake with these biomarkers have been reported previously^43–45^, therefore, for the follow-up mediation analyses in order to evaluate how much MED-mortality risk can be explained by different factors, we also included these factors (**Table 1 and Table 2**).

**Table 2.**
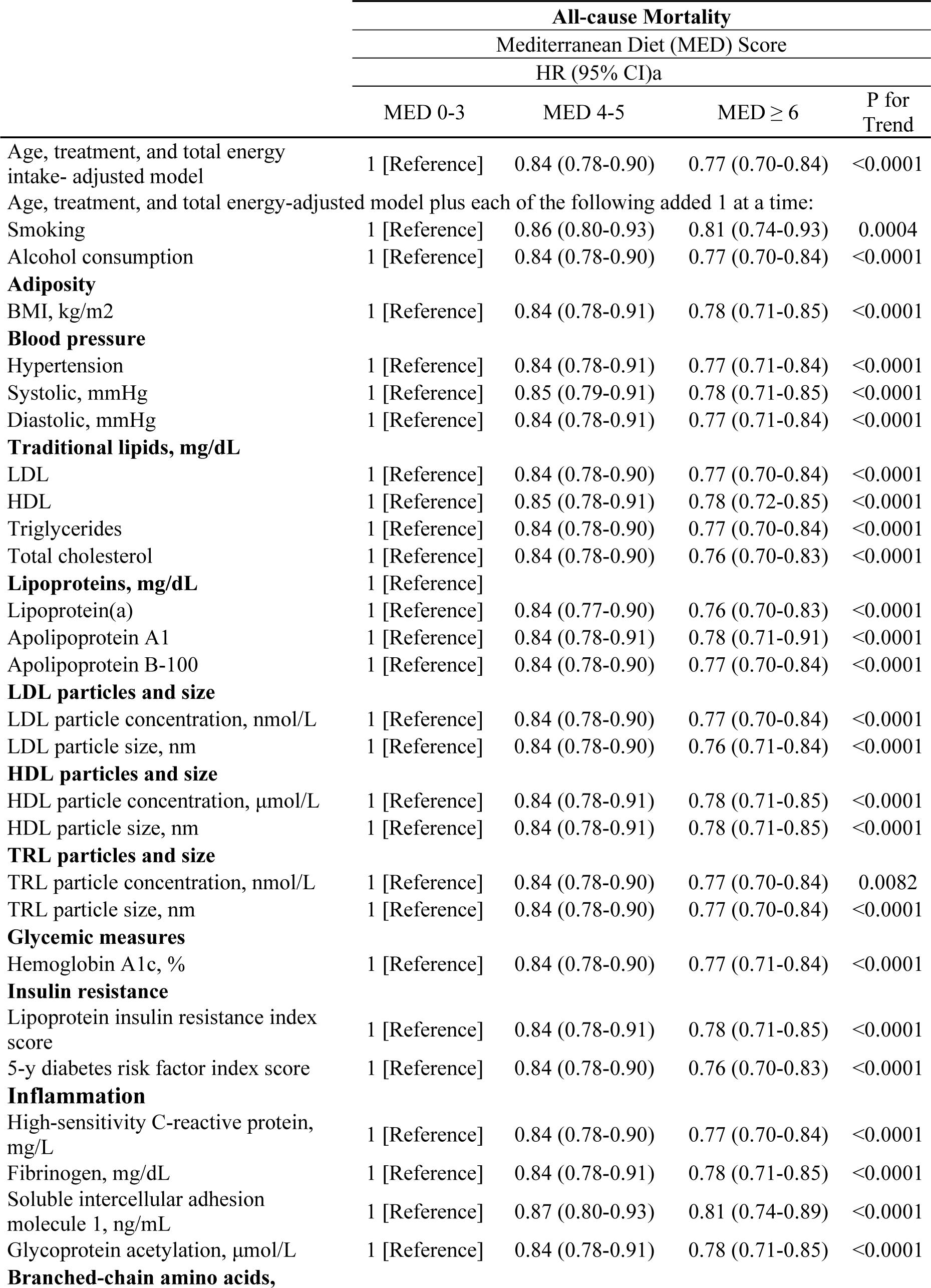
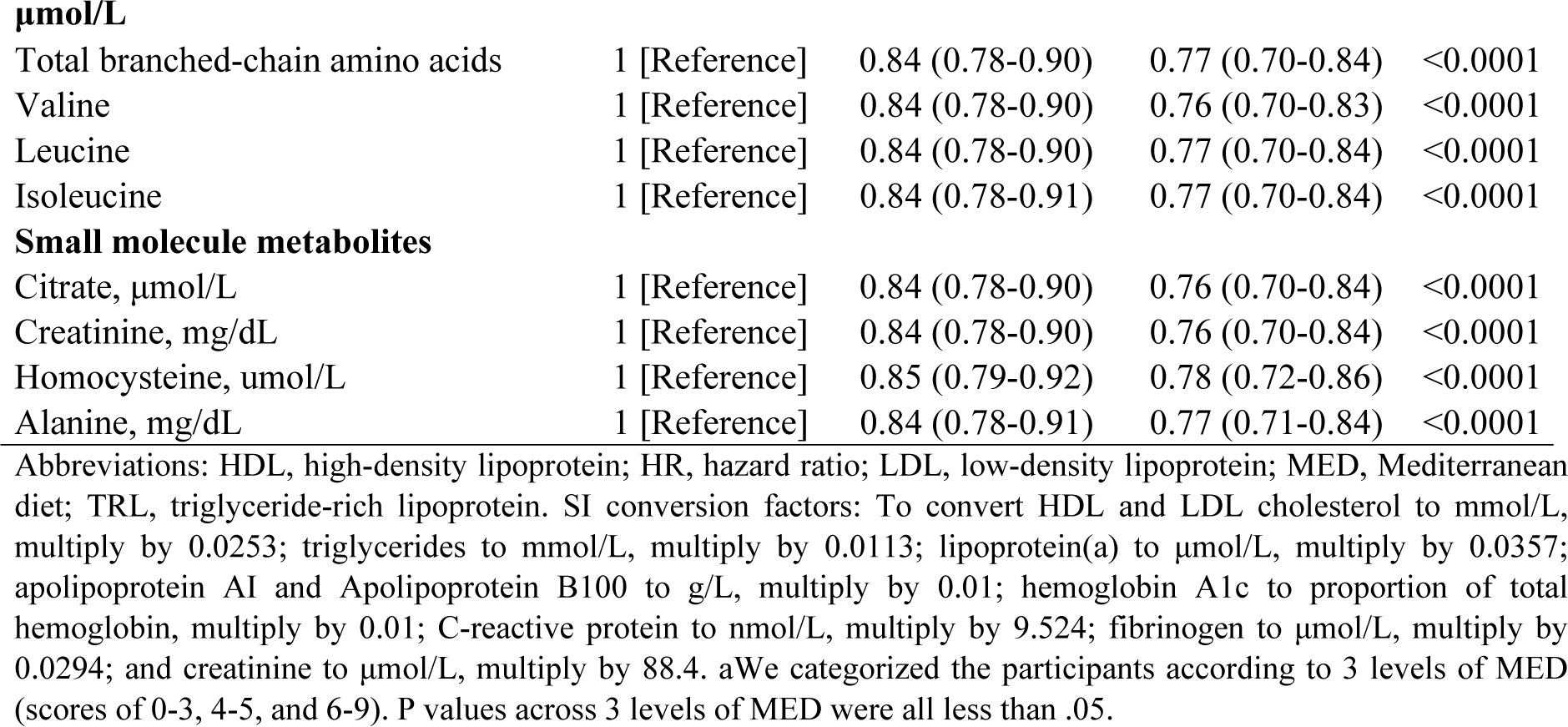
Association of Mediterranean diet intake with all-cause mortality before and after adjustment for risk factors and cardiometabolic biomarkers of risk.

### Mediterranean Diet Intake and Lower Risk of Mortality

During a mean of 24.7 (SD: 4.8) years of follow-up, a total of 3,879 all-cause mortality events occurred, including 935 CVD deaths. The risk of all-cause and CVD mortality decreased linearly (P for linear trend <0.0001) with higher MED intake (**Table S1**). Cumulative incidence curves of the three levels of MED intake with all-cause and CVD mortality risk are shown in **Figure 1** **and** **Figure 2, respectively**. Compared with the reference group (MED intake of 0-3, approximately bottom tertile) and adjusting for age, treatment and energy intake, the relative risk reduction of all-cause mortality associated with the MED intake of 4-5 and ≥ 6 groups were 0.84 (95% CI, 0.78-0.90) and 0.77 (95% CI, 0.70-0.84), respectively (*P* for trend < .0001) (**Table 3**). Similarly, higher MED intake was associated with reduced risk of CVD mortality with respective HRs of 0.86 (95% CI, 0.74-1.01) and 0.83 (95% CI, 0.69-0.99) (P for trend = 0.034), and non-CVD mortality with respective HRs of 0.83 (95% CI, 0.76-0.90) and 0.75 (95% CI, 0.68-0.83) (P for trend = <0.0001) (**Table 3**). Further adjusting for smoking, physical activity, alcohol intake and menopausal factors attenuated the risk reductions somewhat but they remained significant with respective HRs of 0.92 (95% CI 0.85-0.99) and 0.89 (95% CI 0.82-0.98), p for trend 0.0011, for the middle and top MED intake groups with all-cause mortality. Risk reductions were generally similar for CVD mortality, with respective HRs of 0.95 (95% CI 0.81-1.11) and 0.96 (95% CI 0.80-1.16), p for trend 0.6555 for the middle and top MED intake groups with CVD mortality. Risk reductions were also generally similar for non-CVD mortality, with respective HRs of 0.91 (0.83-0.99) and 0.87 (0.79-0.97), p for trend 0.0082 for the middle and top MED intake groups with non-CVD mortality.

**Figure 1.**
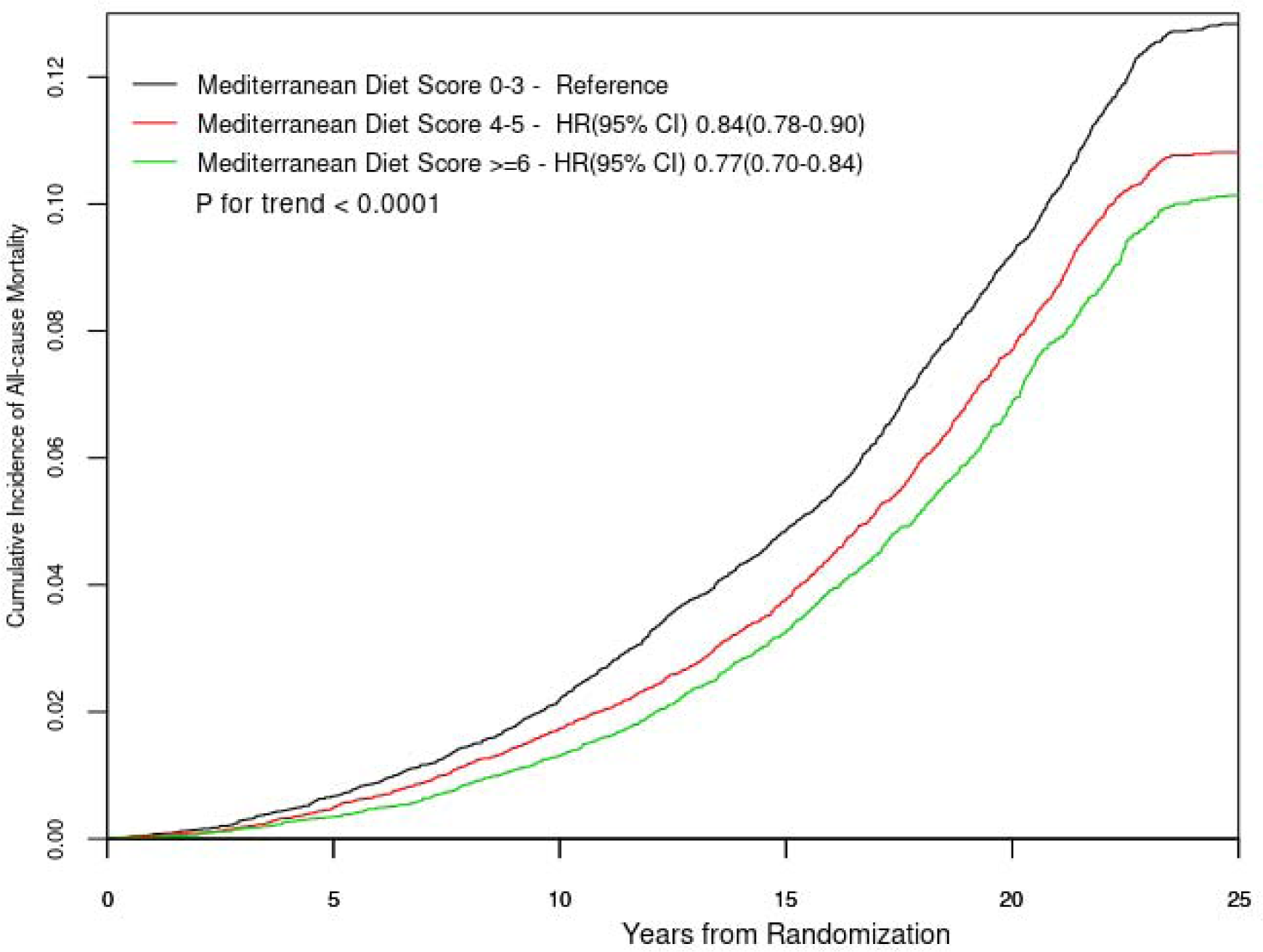
Cumulative survival for the Mediterranean diet in all-cause mortality confirmed person years. The analyses were adjusted for age, treatment, and total energy intake.

**Figure 2.**
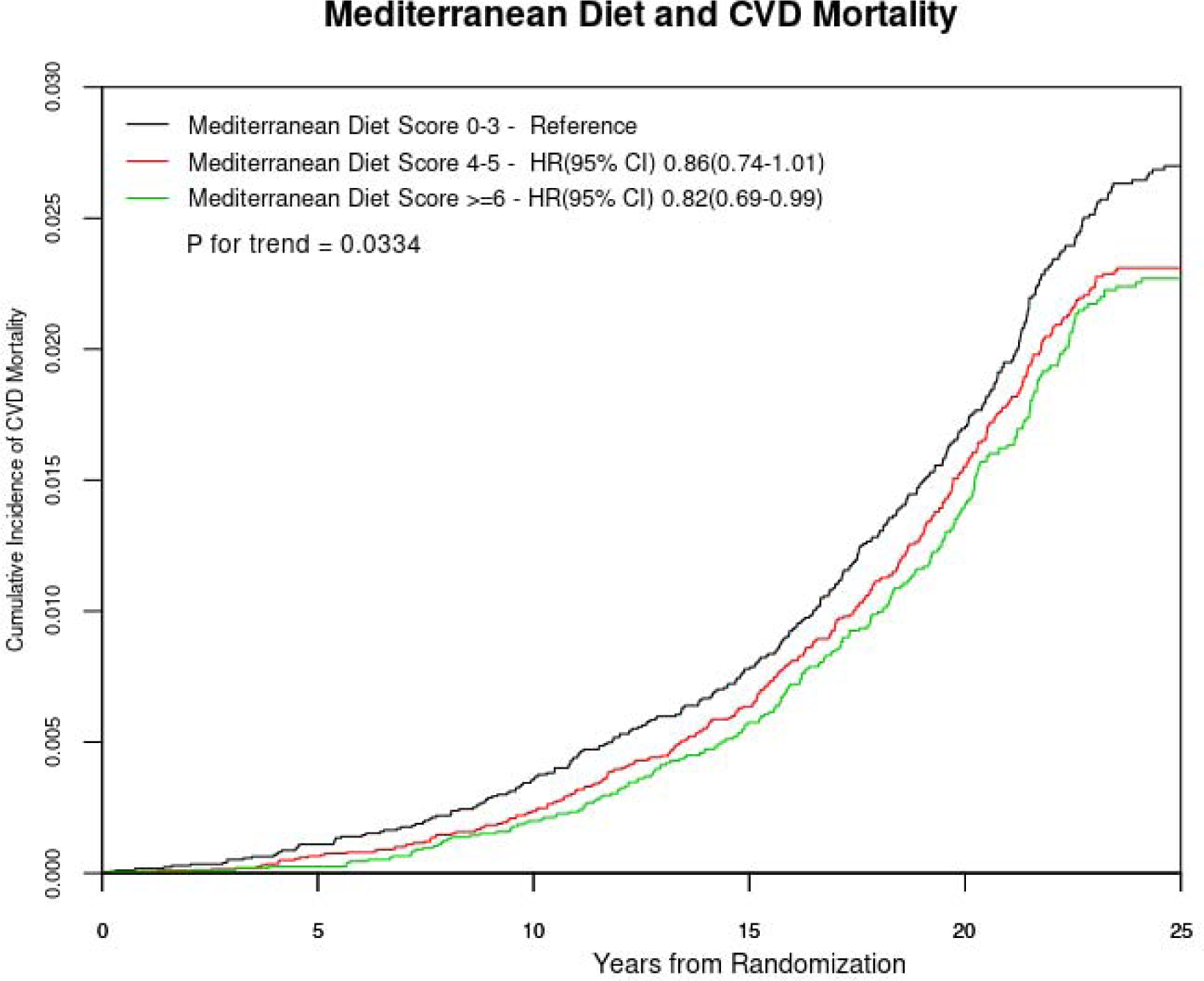
Cumulative survival for the Mediterranean diet in total CVD mortality confirmed person years. The analyses were adjusted for age, treatment, and total energy intake.

**Table 3.** Association of Mediterranean diet with all-cause mortality events before and after adjustment for sets of potential mediators.

|  | Mediterranean Diet Score |  |  | P for Trend |
| --- | --- | --- | --- | --- |
| | MED 0-3 | MED 4-5 | MED $\geq 6$ | |
| Age, treatment, and total energy intake- adjusted model | 1 [Reference] | 0.84 (0.78-0.90) | 0.77 (0.70-0.84) | <0.0001 |
| Basic model* | 1 [Reference] | 0.92 (0.85-0.99) | 0.89 (0.82-0.98) | 0.0011 |
| Basic model plus each set of risk factors below, added 1 group at a time† |  |  |  |  |
| Adiposity | 1 [Reference] | 0.92 (0.85-1.00) | 0.90 (0.82-0.99) | 0.022 |
| Hypertension | 1 [Reference] | 0.92 (0.86-1.00) | 0.90 (0.82-0.98) | 0.0014 |
| Apolipoproteins: lipoprotein(a), apolipoprotein AI, apolipoprotein B100 | 1 [Reference] | 0.92 (0.85-0.99) | 0.89 (0.81-0.98) | 0.011 |
| LDL measures: LDL particle size and concentration, LDL cholesterol, apolipoprotein B100 | 1 [Reference] | 0.92 (0.85-0.99) | 0.90 (0.82-0.98) | 0.013 |
| HDL measure: HDL particle size and concentration, HDL cholesterol, apolipoprotein AI | 1 [Reference] | 0.92 (0.85-0.99) | 0.90 (0.82-0.98) | 0.014 |
| TRL measures: triglyceride-rich lipoprotein particle size and concentrations, triglycerides | 1 [Reference] | 0.92 (0.86-1.00) | 0.90 (0.82-0.99) | 0.022 |
| Hemoglobin A1c | 1 [Reference] | 0.92 (0.85-0.99) | 0.89 (0.82-0.98) | 0.012 |
| Insulin resistance: Lipoprotein insulin resistance index score, 5-y diabetes risk factor index score | 1 [Reference] | 0.92 (0.86-1.00) | 0.90 (0.82-0.99) | 0.018 |
| Inflammation: hsCRP, fibrinogen, sICAM-1, glycoprotein acetylation | 1 [Reference] | 0.93 (0.86-1.00) | 0.91 (0.83-0.99) | 0.028 |
| Branched-chain amino acids | 1 [Reference] | 0.92 (0.85-0.99) | 0.90 (0.82-0.98) | 0.014 |
| Small-molecule metabolites: citrate, creatinine, homocysteine, alaline | 1 [Reference] | 0.93 (0.86-1.00) | 0.91 (0.83-1.00) | 0.032 |
| All of the above | 1 [Reference] | 0.93 (0.86-1.01) | 0.92 (0.83-1.00) | 0.048 |

\*Basic Model included age, randomized treat assignment; total energy intake (TEI, quintiles), smoking, menopausal status, postmenopausal hormone use, physical activity. Participants were followed-up upto 25 years from baseline. †Models were adjusted for the variables in the basic model plus each of the sets of risk factors added 1 group at a time to separate models. ‡Model included variables in the basic model, plus all sets of risk factors included simultaneously in 1 model. Adiposity here refers to BMI.

Per unit increment in MED intake (adjusted for age, treatment, and energy intake), the HRs were 0.94, 95% CI 0.92-0.95, *P*-value < 0.0001 for all-cause mortality, and 0.95, 95% CI 0.92-0.99, *P*-value = 0.015, for CVD mortality. Further adjusting for smoking, physical activity, alcohol intake and menopausal factors resulted in HRs of 0.97, 95% CI 0.95-0.99, *P*-value = 0.0023 for all-cause mortality and 0.99, 95% CI 0.95-1.03, *P*-value = 0.57 for CVD mortality.

In the follow-up analyses, separate Cox regression models were adjusted for each risk factor or biomarker one at a time in addition to age, treatment and energy intake (**Table 2**). We observed some attenuation of all-cause mortality associations after adjusting for risk factors/biomarkers one at a time, although most associations remained significant.

Next, in order to better characterize the extent to which the reduced risk of all-cause mortality associated with MED intake was influenced by potential sets of mediators from different physiological pathways, we grouped the risk factors/biomarkers into physiological sets and each set of mediators was added at a time to the basic model (**Table 3 and** **Figure 3**). We computed the degree of the mediation association for MED diet with all-cause mortality which could be attributed to each set of potential mediators. For MED intake and all-cause mortality, we observed small-molecule metabolites (e.g., citrate, (e.g., homocysteine, alanine) and inflammation explained the largest contributions to lower risk of overall mortality (14.8% and 13.0% respectively), with lesser contributions from TRLs (10.2%), adiposity (10.2%), and insulin resistance (7.4%). Smaller contributions (<3%) were seen for HDL or LDL measures, hypertension, BCAAs, and HbA1c. When all the risk factors/biomarkers were added together in the regression model, we observed 21.3% total mediation effect.

**Figure 3.**
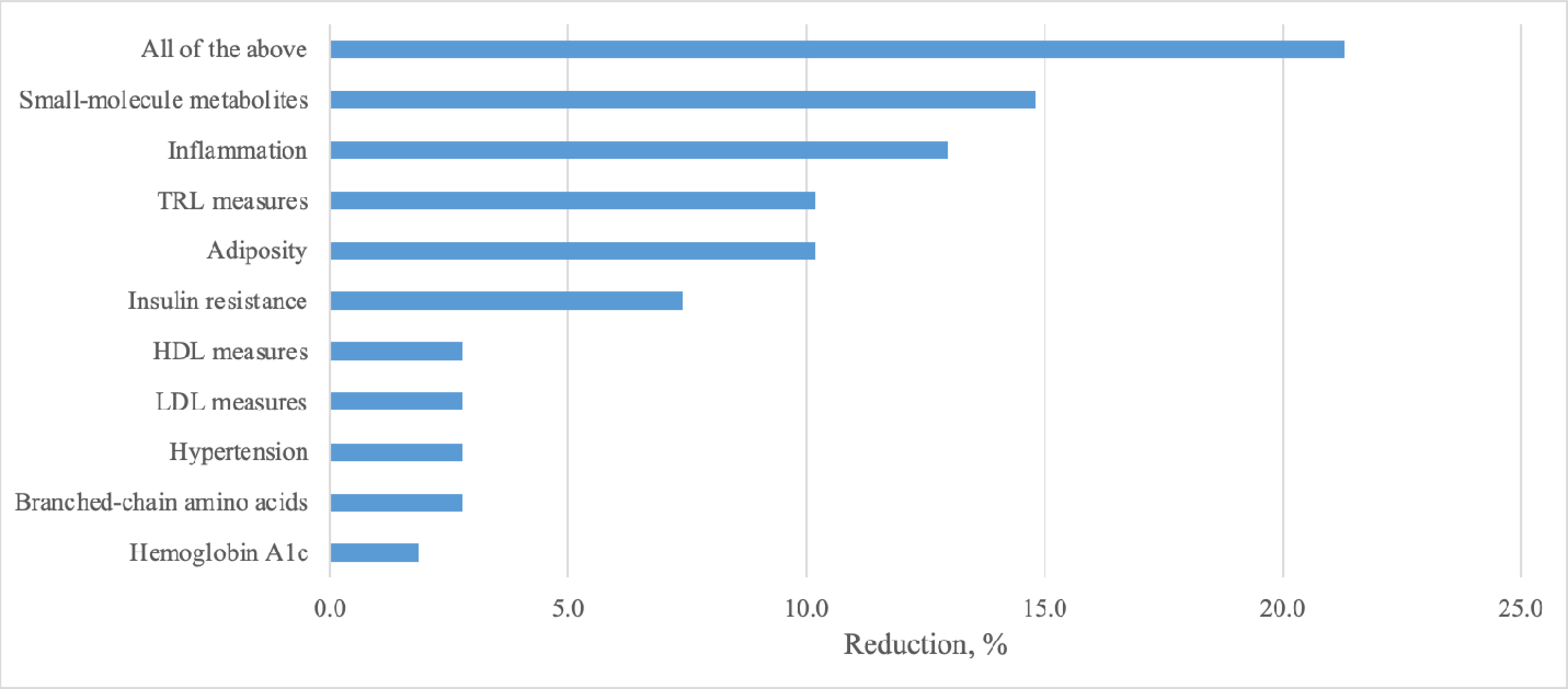
The proportion of all-cause mortality reduction for a Mediterranean diet score of 6 or greater (vs the reference group of 0-3) is shown for potential groups of mediators, calculated as (HR _basic_ model –HR _adjusted model_)/ (HR _basic model_ -1) *100. Apolipoproteins B, AI and Lpa did not contribute in mediating the relationship of MED intake with all-cause mortality. The basic model included age, randomized treat assignment, energy intake, smoking, alcohol intake, menopausal status, postmenopausal hormone use, and physical activity.

In sensitivity analyses, we compared the reported mediation results for single biomarkers through both the counterfactual framework approach^40^ as well as the standard mediation approach, and the results were generally similar across both approaches (**Table S2 and Figure 4**).

## DISCUSSION

In this large-scale prospective study of 25,315 initially healthy US women who were followed for 25 years, we observed that higher consumption of MED intake was associated with nearly one fifth reduction in all-cause mortality, CVD and non-CVD mortality among women. Furthermore, the MED associated all-cause mortality risk reduction was explained partially by potential mediation through small molecules metabolites (e.g. alanine), inflammatory biomarkers, TRL measures, adiposity and insulin resistance and to a much lesser extent by blood pressure, HDL, LDL, apolipoprotein(B), lipoprotein(a) or glycemic measures.

Our findings of lower risk of long-term all-cause and CVD mortality among women with higher consumption of MED are consistent with the data from prior studies in US populations, which reported that higher MED consumption was associated with 16% reductions in all-cause and CVD mortality^13^, and other cohorts based on the US and non US population have reported beneficial effects of MED consumption^12, 18, 19, 46^. The study findings with long-term mortality in this population of women is also consistent with our prior study evaluating risk of CVD events, which found one quarter reduction in total CVD events (fatal and non-fatal) over a 12-year period for the top vs bottom MED intake groups^31^.

Prior shorter-term studies have also demonstrated beneficial effects of MED consumption in relation to cardiometabolic, inflammatory and lipid biomarkers. In a cross-sectional study, adherence to MED was associated with lower levels of CRP and interleukin-6 and improved endothelial function^24–26^. In a 2-years intervention trial of MED intake was associated with reduced serum concentrations of CRP, interleukin -6 (IL-6), IL-7, and IL-18 as well as decreased insulin resistance^47^. These prior studies are consistent with the current findings in particular of mediation of association for inflammatory and insulin resistance biomarkers. In most prior studies, MED diet did not result in substantial changes in total cholesterol, LDL cholesterol, or lipoprotein(a), consistent with the current results. In a 3-months dietary clinical trial, MED consumption was better in reducing oxidized LDL levels in comparison to a low-fat diet^48^. It is possible that more functional assessments of LDL and HDL may be related to MED diet intake or benefit.

The current study is a large-scale prospective epidemiological study of validated dietary measures, detailed and comprehensive measures of traditional and novel NMR based biomarkers as well as large number of incident mortality cases during 24 years of follow-up of US women. Nonetheless, there are several potential limitations. Study participants were of middle aged well-educated female health professional who were predominantly of non-Hispanic White ancestry. Dietary intake was assessed through FFQs and we cannot rule out the possibility of exposure misclassification, under- and overreporting which might attenuate MED diet and mortality associations towards the null. Both dietary assessment as well as blood biomarkers were assessed at baseline. The potential residual confounding from unmeasured variables cannot be ruled out, however, the association remained significant after adjustment for potential confounders. It is possible that some of the covariates, such as hypertension and adiposity, could be influenced by the MED intake which suggests these variables could be confounders and/or mediators.

In summary, in this large-scale study of initially healthy US women there was approximately one fifth reduction in long-term all-cause mortality with MED diet. Our results suggest that a proportion of the lower risk of mortality may be accounted for by cardiometabolic risk factors in particular for biomarkers related to metabolism, inflammation, insulin resistance, TRL pathways and adiposity but not those related to total, LDL cholesterol, lipoprotein(a) or standard glycemic measures such as HbA1c. Despite this, most of the potential benefit of the MED intake and morality remains unexplained and future studies should examine other pathways that could potentially mediate the MED-associated lower mortality.

## Data Availability

The authors had full access to and take full responsibility for the integrity of the data. All authors have read and agree to the manuscript as written.

## ACKNOWLEDGEMENT

The Women’s Health Study is supported by the National Institutes of Health (grants CA047988, HL043851, HL080467, HL099355, and UM1 CA182913). Dr Ahmad was supported through a career starting research grants from Swedish Research Council (2022-01460) and FORMAS (2020-00989) and also research grant from the EpiHealth, Sweden. Dr Demler was supported by a K award from the National Heart, Lung, and Blood Institute of the National Institutes of Health under award number K01HL135342-02. Dr Mora was supported by the research grants from the National Institute of Diabetes and Digestive and Kidney Diseases (grant DK112940); National Heart, Lung, and Blood Institute (grants R01HL134811, R01HL117861 and K24 HL136852); American Heart Association (grant 0670007N); and the Molino Family Trust. In addition, LabCorp provided the LipoProfile IV results to the study at no additional cost.

## Disclosures

Dr. Mora is listed as co-inventor on a patent regarding the use of an NMR glycosylation biomarker (GlycA) in relation to colorectal cancer risk. All other coauthors have no conflict of interests.

## Notes

### Author Declarations

All study participants provided written consent and the study was approved by the institutional review board of Brigham and Women's Hospital, Boston, Massachusetts.

